## Additional detail on materials and methods for "Quantifying epidemiological drivers of *gambiense* human African Trypanosomiasis across the Democratic Republic of Congo"

### Supplementary Information: Quantifying epidemiological drivers of *gambiense* human African trypanosomiasis across the Democratic Republic of Congo. S1 Materials and Methods

Ronald E Crump[1,2,3◕*], Ching-I Huang[1,2◕], Ed Knock[1,4], Simon E F Spencer[1,4], Paul Brown[1,2], Erick Mwamba Miaka[5], Chansy Shampa[5], Matt J Keeling[1,2,3], and Kat S Rock[1,2]

[1]Zeeman Institute for System Biology and Infectious Disease Epidemiology Research, The University of Warwick, Coventry, U.K.
[2]Mathematics Institute, The University of Warwick, Coventry, U.K.
[3]The School of Life Sciences, The University of Warwick, Coventry, U.K.
[4]The Department of Statistics, The University of Warwick, Coventry, U.K.
[5]PNLTHA, Kinshasa, D.R.C.

October 26, 2020

◕These authors contributed equally to this work.

#### S1.1 Data

**HAT Atlas data** The HAT Atlas data for DRC were provided in a spreadsheet format. Records were annually aggregated gHAT case records, aggregated by year, surveillance type and location as defined by multiple fields. There were 117,573 rows in this file; of which 111,408 had an entry in the geolocation (longitude and latitude) fields.

Passive surveillance records with missing or zero case numbers; and active surveillance records with both missing or zero numbers screened and missing or zero case numbers were dropped from the dataset.

This left 111,454 records (105,979 with filled geolocation fields). These records were associated with 23,424 unique combinations of former province, health zone, health area, location and territory identifiers; 20,423 of which had geolocation information and 3,001 did not. We will refer to these 24,424 geographical records as gHAT locations in this document.

Table S1.1: Number of HAT Atlas records with different combinations of former province, health zone and health area recorded.

| Recorded region identifiers: | | | Number |
|---|---|---|---|
| Former province | Health zone | Health area | n |
| ✓ | ✓ | ✓ | 106823 |
| ✓ | ✓ | ✓ | 391 |
| ✓ | ✓ | | 3001 |
| ✓ | | ✓ | 14 |
| ✓ | | | 1225 |

**DRC Shapefile** A recent shapefile for DRC was provided by UCLA (Personal communication). The shapefile contains health zones (an organisational unit with a typical population size around 100,000) across DRC; health areas (nested within health zones, these areas are typically home to around 10,000 people) for the former province of Bandundu and part of Equateur and Haut Lomami, and post–2015 province identifiers. Former province was added to these records (post–2015 provinces being nested within former province).

**Additional geographic information** The following geographical information was obtained from the Humanitarian Data Exchange [S2]:

- a health zone shapefile from the United Nations Office for the Coordination of Humanitarian Affairs (OCHA);
- an OCHA file of geolocations of localities; and
- a file of geolocations of health facilities from the Global Healthsite Mapping Project.

These data were used to assist in matching and locating the gHAT data, by providing alternative spellings of names and potentially geolocations for non-geolocated gHAT locations. The locality and health facility lists were concatenated, and this enlarged locality set and the OCHA health zone map were assigned geographical identifiers as per our shapefile of choice.

**Matching HAT Atlas records to DRC shapefile** Geographical identifiers associated with the HAT Atlas/gHAT location and geographical data were sanitised to assist with matching. This involved removing diacritical marks, conversion to lowercase, collapsing whitespace within identifiers to a single space, removal of leading and trailing whitespace, converting from roman to arabic numerals, removing leading m, n or g from words where they were followed by a consonant, removing leading t from words when followed by an s, and collapsing words into a single string. In addition some specific manual edits were performed during the process as they became apparent.

Matching was then applied sequentially to the gHAT locations; such that once a match had been achieved for any given gHAT location it did not act as input to subsequent steps.

1. gHAT locations with known former province (FP), health zone (HZ), health area (HA) and geolocation were located on the UCLA and OCHA shapefiles. If the FP, HZ and HA matched the values for either the UCLA or OCHA shapefiles at that point, a match was judged to have occurred and the geolocation was accepted. This matched 3,579 of the gHAT locations.
2. gHAT locations with known former province (FP), health zone (HZ) and geolocation were located on the UCLA and OCHA shapefiles. If the FP and HZ matched the values for either the UCLA or OCHA shapefiles at that point, a match was judged to have occurred and the geolocation was accepted. This matched 13,413 of the gHAT locations. *The 16,992 gHAT locations matched in these two steps accounted for 97,520 of the HAT records (87.5%).*
3. Where a gHAT location was associated with a recent active screening event (defined as an active screening record in or after 2012 with a number screened greater than 10), the geolocation was accepted. *This matched a further 454 of the gHAT locations to give a total of 100,747 gHAT records matched (90.4%).*
4. Matching to the locality information:
   (a) if health zone and location identifier match, the locality's geolocation was assigned to the gHAT location. *Matching 44 gHAT locations, giving a total of 100,811 gHAT records matched (90.5%).*
   (b) if former province and location identifier match, the locality's geolocation was assigned to the gHAT location. *This matched 331 gHAT locations, giving a total of 101,508 gHAT records matched (91.1%).*
5. Matching to shapefile by geographic identifiers only:
   (a) Former province, health zone and health area all match. *Matched 523 gHAT locations, giving a total of 102,349 gHAT records matched (91.8%).*
   (b) Former province and health zone match. *Matched 3636 gHAT locations, giving a total of 110,021 gHAT records matched (98.7%).*
   (c) Former province and OCHA shapefile health zone name match. *Matched 74 gHAT locations, giving a total of 110,126 gHAT records matched (98.8%).*

(d) Former province and health area match. *Matched 60 gHAT locations, giving a total of 110,195 gHAT records matched (98.9%).*

(e) Health zone name match. *Matched 10 gHAT locations, giving a total of 110,210 gHAT records matched (98.9%).*

#### S1.2 Modelling passive detection and its improvement

There are two sources of passive detection improvements considered in our model: a rapid improvement due to the introduction of the card agglutination test for trypanosomes (CATT) test in all health zones in 1998 and a gradual improvement over time in the former Bandundu and Bas Congo provinces around 2008 and mid-2015 respectively. Prior distributions and percentiles of parameters related to passive detection and it's improvement over time are summarised in Table S1.2.

Improvement in passive surveillance systems over time is considered across the whole of Bandundu and Bas Congo. The province level staging data in Bandundu suggested that passive surveillance systems in Bandundu have improved over time, which was confirmed by PNLTHA and is also supported by previous modelling work [S1]. In Bas Congo, FIND implemented the use of rapid diagnostic tests (RDT) from 2015. Staging information which was available at the province–level from 2000–2012 from the paper of Lumbala *et al* [S3], and in the HAT Atlas data for 2015 and 2016. To inform the health zone-level analyses, a province level fit was carried out to the staged case data of Lumbala *et al.* augmented with the HAT Atlas data aggregated to the former province level for the years 2013-2016. These analyses provided no evidence for improvement in passive surveillance systems (in line with the simple sigmoidal model assumed) for any provinces other than Bandundu and Bas Congo. For Bandundu and Bas Congo, gamma distributions were fitted to the province level posterior samples of $\eta_{H_{\text{amp}}}$, $\gamma_{H_{\text{amp}}}$ and $d_{\text{steep}}$. The shape ($k$) and scale ($\theta$) for these fitted distributions were used as the parameters of gamma prior distributions of $\eta_{H_{\text{amp}}}$, $\gamma_{H_{\text{amp}}}$ and $d_{\text{steep}}$ in all health zones of Bandundu and Bas Congo. In Bandundu health zones a scaled and shifted beta distribution was used as the prior for $d_{\text{change}}$. The use of a broader prior for $d_{\text{change}}$ than would have resulted from using the province–level posterior distribution resulted from comparing aggregate health zone-level results with province-level observed data. In Bas Congo health zones a fixed value of 2015.5 was used for $d_{\text{change}}$.

Priors for the health zone-level $\eta_H^{\text{post}}$ and $\gamma_H^{\text{post}}$ parameters were also informed by the province-level fits. Gamma prior distributions were used which had the same mode $(\text{mode} = (k-1)\,\theta$ as Gamma distributions fitted to the province-level posterior distribution of the parameters; and a standard deviation $(s.d. = \sqrt{k}\theta)$ of $3 \times 10^{-3}$ for $\eta_H^{\text{post}}$ and $1 \times 10^{-4}$ for $\gamma_H^{\text{post}}$, being arbitrarily selected higher variation than the province-level posterior distributions.

#### S1.3 Modelling vector control

Fig. S1.1 shows the dynamics of tsetse populations (under the simple model) where targets are either moderately effective, with a 60% reduction (lower than in Guinea), or highly effective, with a 90% population density reduction in a year (as seen in Uganda).

The function which describes the probability of both hitting a target and dying is time dependent (days) from when the targets where placed:

$$f_T(t) = f_{\text{max}} \left(1 - \frac{1}{1 + \exp(-0.068(\text{mod}(t, 182.5) - 127.75))}\right) \tag{S1.3.1}$$

and $f_{\text{max}}$ is chosen such that the tsetse population after one year is at the observed/assumed percentage reduction. For the simplified model this is given by $f_{\text{max}} = 0.0305$ for a 60% reduction and $f_{\text{max}} = 0.0750$ for a 90% reduction.

#### S1.4 Imputation of missing numbers screened information

There are instances in the data where the number of cases from active screening within year $t$ ($A_D(t) = A_{D1} + A_{D2}$) is not consistent with the number of people recorded as having been screened in that year for that health zone ($z(t)$), i.e. $A_D(t) > z(t)$. In this situation, if $A_D(t) < 20$ we assume that these people have attended a screening outside their home health zone and the record has been correclty allocated to their

Table S1.2: **Parameterisation of passive detection improvement.** Notation and brief description of fitted parameters related to passive detection improvement plus their within former province prior distributions and [2.5th, 50th & 97.5th] percentile.

| Parameter | | | |
|---|---|---|---|
| | **Province** | **Prior distribution** | **Percentiles of prior distribution** |
| $\eta_H^{\text{post}}$ – Treatment rate from stage 1, 1998 onwards | | | |
| | **Bandundu** | $\Gamma\big(3.54, 5.32 \times 10^{-5}\big)$ | [4.59, 17.1, 42.9]$\times 10^{-5}$ |
| | **Bas Congo** | $\Gamma\big(12.0, 2.89 \times 10^{-5}\big)$ | [1.78, 3.36, 5.68]$\times 10^{-4}$ |
| | **Equateur** | $\Gamma\big(4.92, 4.51 \times 10^{-5}\big)$ | [7.12, 20.7, 45.7]$\times 10^{-5}$ |
| | **Kasai Occidental** | $\Gamma\big(10.9, 3.03 \times 10^{-5}\big)$ | [1.64, 3.20, 5.53]$\times 10^{-4}$ |
| | **Kasai Oriental** | $\Gamma\big(2.90, 5.87 \times 10^{-5}\big)$ | [3.38, 15.1, 41.5]$\times 10^{-5}$ |
| | **Katanga** | $\Gamma\big(1.29, 8.79 \times 10^{-5}\big)$ | [5.88, 86.2, 376]$\times 10^{-6}$ |
| | **Kinshasa** | $\Gamma\big(1.26, 8.91 \times 10^{-5}\big)$ | [5.44, 84.4, 376]$\times 10^{-6}$ |
| | **Maniema** | $\Gamma\big(4.25, 4.85 \times 10^{-5}\big)$ | [5.90, 19.0, 44.3]$\times 10^{-5}$ |
| | **Orientale** | $\Gamma\big(1.16, 9.27 \times 10^{-5}\big)$ | [4.24, 79.0, 373]$\times 10^{-6}$ |
| $\gamma_H^{\text{post}}$ – Treatment rate from stage 2, 1998 onwards | | | |
| | **Bandundu** | $\Gamma\big(2.45, 1.92 \times 10^{-3}\big)$ | [7.59, 40.7, 121]$\times 10^{-4}$ |
| | **Bas Congo** | $\Gamma\big(1.48, 2.47 \times 10^{-3}\big)$ | [2.54, 28.7, 114]$\times 10^{-4}$ |
| | **Equateur** | $\Gamma\big(1.95, 2.15 \times 10^{-3}\big)$ | [4.88, 35.0, 118]$\times 10^{-4}$ |
| | **Kasai Occidental** | $\Gamma\big(1.71, 2.29 \times 10^{-3}\big)$ | [3.65, 31.9, 116]$\times 10^{-4}$ |
| | **Kasai Oriental** | $\Gamma\big(1.49, 2.46 \times 10^{-3}\big)$ | [2.60, 28.8, 114]$\times 10^{-4}$ |
| | **Katanga** | $\Gamma\big(1.53, 2.43 \times 10^{-3}\big)$ | [2.78, 29.4, 115]$\times 10^{-4}$ |
| | **Kinshasa** | $\Gamma\big(1.68, 2.31 \times 10^{-3}\big)$ | [3.50, 31.5, 116]$\times 10^{-4}$ |
| | **Maniema** | $\Gamma\big(2.60, 1.86 \times 10^{-3}\big)$ | [8.45, 42.3, 122]$\times 10^{-4}$ |
| | **Orientale** | $\Gamma\big(2.54, 1.88 \times 10^{-3}\big)$ | [8.11, 41.7, 122]$\times 10^{-4}$ |
| $\eta_{H_{\text{amp}}}$ – Relative improvement in passive stage 1 detection rate | | | |
| | **Bandundu** | $\Gamma(2.01, 1.05)$ | [0.258, 1.77, 5.87] |
| | **Bas Congo** | $\Gamma(5.23, 1.70)$ | [2.98, 8.33, 18.0] |
| $\gamma_{H_{\text{amp}}}$ – Relative improvement in passive stage 2 detection rate | | | |
| | **Bandundu** | $\Gamma(1.001, 5)$ | [0.127, 3.47, 18.5] |
| | **Bas Congo** | $\Gamma(1.46, 1.26)$ | [0.126, 1.45, 5.81] |
| $d_{\text{steep}}$ – Speed of improvement in passive detection rate | | | |
| | **Bandundu** | $\Gamma\big(39.6, 2.70 \times 10^{-2}\big)$ | [0.761, 1.06, 1.42] |
| | **Bas Congo** | $\Gamma(3.21, 1.45)$ | [1.03, 4.18, 10.9] |

home health zone and we set $z(t) = A_D(t)$. However, where $A_D(t) \geq 20$ we assume that a screening must have taken place in the health zone but that only the positive test results have been recorded. To allow use of these records we impute the missing negative test results $(A_D^-(t))$ during the MCMC analysis.

We use a hierarchical prior conditional on the other model parameters ($\theta$) for $A_D^-(t)$; $A_D^-(t)|\theta \sim \text{NB}(A_D(t), P^+(\theta))$, where $P^+(\theta)$ is the probability of a positive active screening result given the current $\theta$ (obtained from solutions of the ODE up to this point in time). The proposal distribution for $A_D^-(t)$ is the same as the conditional prior, and these cancel from the posterior probability. The value $\hat{z}(t) = A_{D1} + A_{D2} + A_D^-(t)$ is used in place of the unknown $z(t)$ in the likelihood calculation.

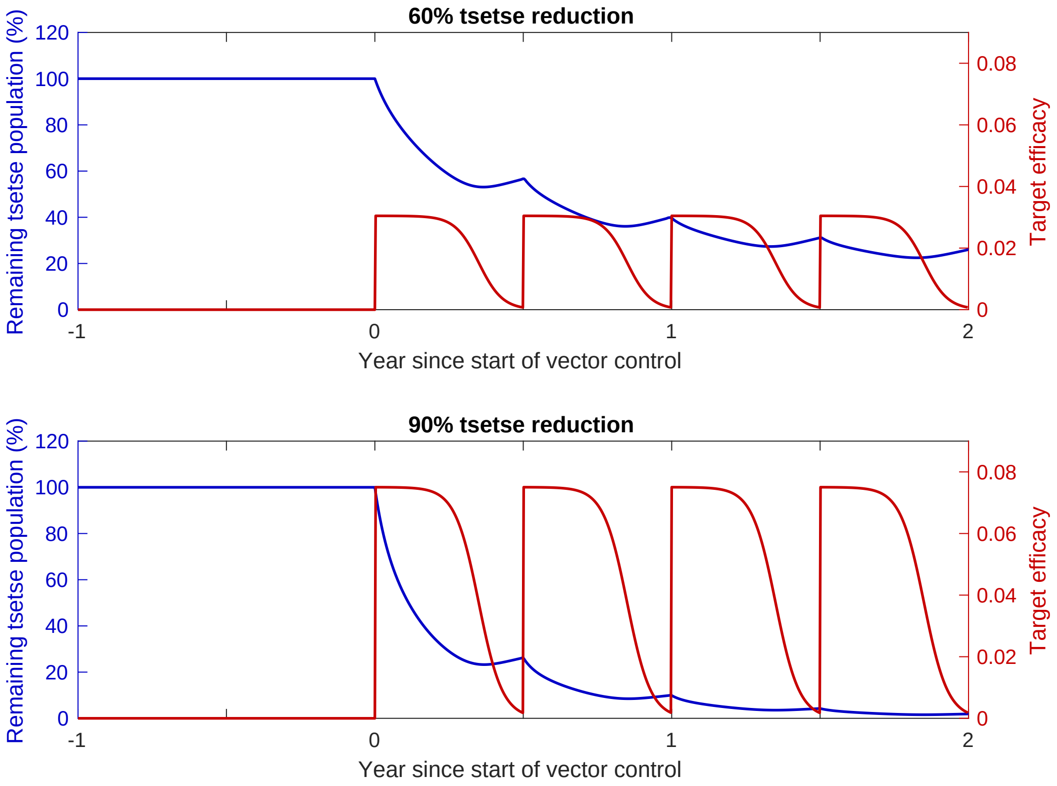


Figure S1.1: **Impact of tiny targets on tsetse density.** The figures show the how varying target efficacy (red line) impacts tsetse population density (blue line). Target efficacy is measured as the proportion of a host-seeking tsetse which will both hit the tiny target and die as a result. The graphs, reproduced from [S4], show the necessary efficacy of targets needed to reduce density by 60% (top) and 90% (bottom) by the end of the first year.

Table S1.3: **Imputation of negative active screening results.** Former province, health zone, year and active case numbers ($A_D(t)$) where imputation of $A_D^-(t)$ was performed; plus median and 95% credible intervals of posterior distributions of $A_D^-(t)$.

| Former province | Health zone | Year ($t$) | $A_D(t)$ | $A_D^-(t)$ | |
|---|---|---|---|---|---|
| | | | | Median | [95% CI] |
| Kinshasa | Maluku 2 | 2000 | 30 | 6052 | [4020, 9101] |
| | | 2001 | 102 | 19653 | [13792, 28992] |
| | Mont Ngafula 1 | 2000 | 303 | 214019 | [125975, 277536] |
| | Nsele | 2001 | 147 | 98902 | [70301, 149644] |
| Orientale | Doruma | 2007 | 320 | 17996 | [15550, 20466] |
| | | 2008 | 152 | 13668 | [10906, 16676] |
| | | 2011 | 215 | 40324 | [29970, 48452] |
