## Supplementary material for "Quantifying epidemiological drivers of *gambiense* human African Trypanosomiasis across the Democratic Republic of Congo": Figures and tables based on posterior distributions of fitted parameters

### Supplementary Information: Quantifying epidemiological drivers of *gambiense* human African trypanosomiasis across the Democratic Republic of Congo. S2 Posteriors of fitted parameters

Ronald E Crump[1,2,3◑*], Ching-I Huang[1,2◑], Ed Knock[1,4], Simon E F Spencer[1,4], Paul Brown[1,2], Erick Mwamba Miaka[5], Chansy Shampa[5], Matt J Keeling[1,2,3], and Kat S Rock[1,2]

[1]Zeeman Institute for System Biology and Infectious Disease Epidemiology Research, The University of Warwick, Coventry, U.K.
[2]Mathematics Institute, The University of Warwick, Coventry, U.K.
[3]The School of Life Sciences, The University of Warwick, Coventry, U.K.
[4]The Department of Statistics, The University of Warwick, Coventry, U.K.
[5]PNLTHA, Kinshasa, D.R.C.

October 26, 2020

◑These authors contributed equally to this work.

#### S2.1 Posterior characteristics for example health zones

Table S2.1 shows summaries of posterior distributions of all fitted parameters in two example health zones: Kwamouth in the former Bandundu province and Tandala in the former Equateur province. Kwamouth is categorised as a high-risk health zone and Tandala is at low-risk. Our fitting results show that Kwamouth has higher $R_0$ and $r$ but lower active screening specificity, reporting rate and passive detection rate than Tandala. These factors provide some possible explanation for why gHAT is more persistent in Kwamouth, as well as having more cases reported every year.

Table S2.1: **Posteriors of fitted parameters.** Notation, brief description, and [2.5th, 50th & 97.5th] percentile of posteriors for fitted parameters.

| **Notation** | **Description** | **Kwamouth** | **Tandala** |
|---|---|---|---|
| $R_0$ | Basic reproduction number (NGM approach) | [1.06, 1.09, 1.15] | [1.006, 1.009, 1.014] |
| $r$ | Relative bites taken on high-risk humans | [3.06, 6.49, 10.98] | [1.30, 2.04, 4.26] |
| $k_1$ | Proportion of low-risk people | [0.82, 0.90, 0.95] | [0.85, 0.95, 0.99] |
| $\eta_H^{\text{post}}$ | Treatment rate from stage 1, 1998 onwards (days$^{-1}$) | [0.60, 1.29, 3.06] $\times 10^{-4}$ | [1.11, 2.74, 4.99]$\times 10^{-4}$ |
| $\gamma_H^{\text{post}}$ | Exit rate from stage 2 (treatment or death), 1998 onwards (days$^{-1}$) | [0.42, 1.79, 4.90] $\times 10^{-3}$ | [1.72, 3.60, 8.98]$\times 10^{-3}$ |
| $b_{\gamma_H^{\text{pre}}}$ | Relative exit rate from stage 2 factor, pre-1998 | [0.73, 0.92, 1.00] | [0.54, 0.69, 0.94] |
| $\gamma_H^{\text{pre}}$ | Exit rate from stage 2 (treatment or death), pre-1998 (days$^{-1}$) | [0.33, 1.64, 4.65] $\times 10^{-3}$ | [1.03, 2.53, 7.14]$\times 10^{-3}$ |
| Spec | Active screening diagnostic specificity | [0.9986, 0.9991, 0.9997] | [0.9997, 0.9998, 0.9999] |
| $u$ | Proportion of stage 2 passive cases reported | [0.18, 0.28, 0.41] | [0.29, 0.39, 0.51] |
| $d_{\text{change}}$ | Midpoint year for passive improvement | [2004.3, 2005.8, 2007.3] | – |
| $\eta_{H_{\text{amp}}}$ | Relative improvement in passive stage 1 detection rate | [0.85, 2.50, 5.74] | – |
| $\gamma_{H_{\text{amp}}}$ | Relative improvement in passive stage 2 detection rate | [0.24, 0.53, 0.99] | – |
| $d_{\text{steep}}$ | Speed of improvement in passive detection rate (years$^{-1}$) | [0.68, 0.94, 1.27] | – |

#### S2.2 Joint posteriors for example health zones

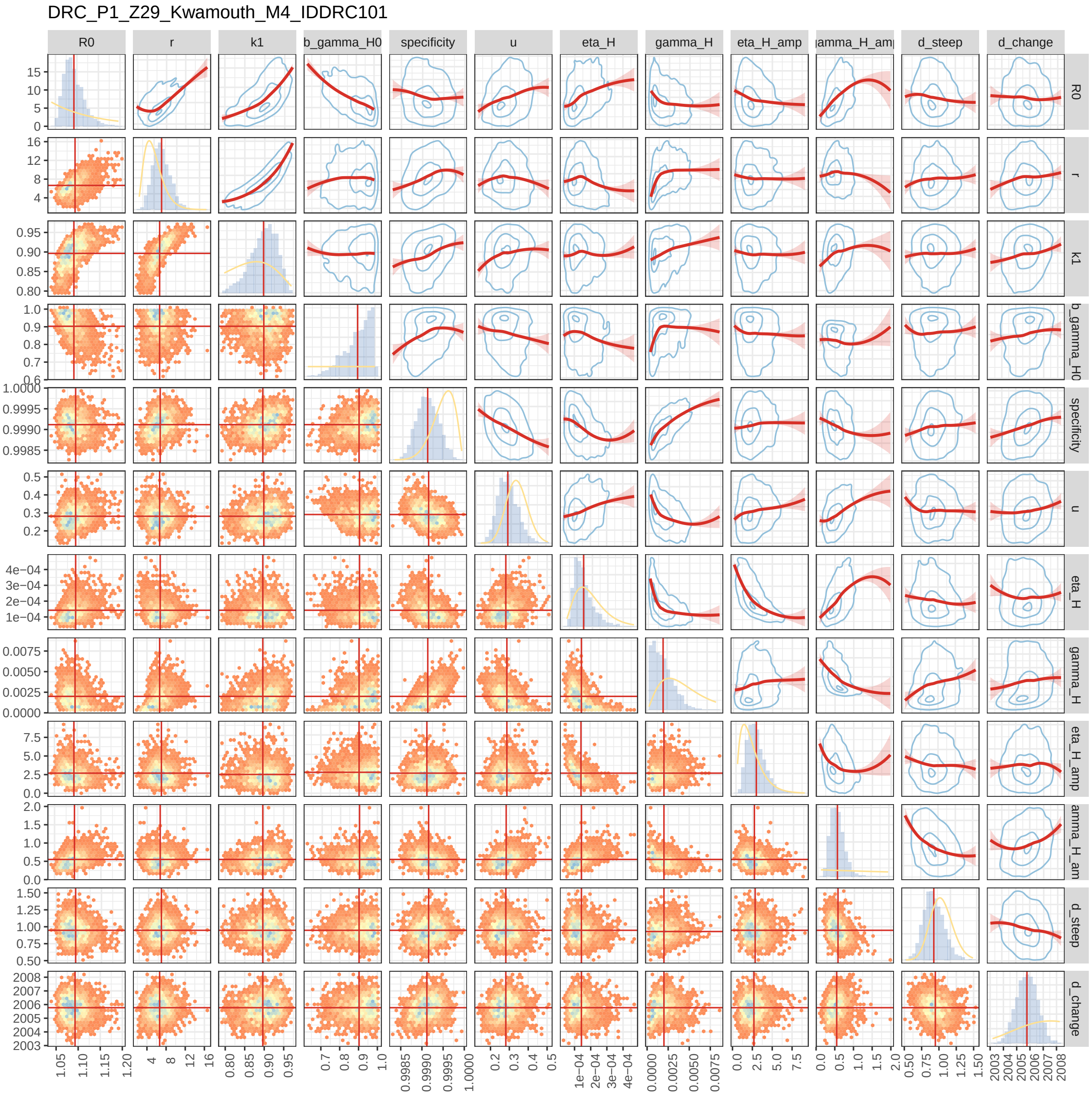


Figure S2.1: Joint posterior distributions for Kwamouth health zone in the former Bandundu province.

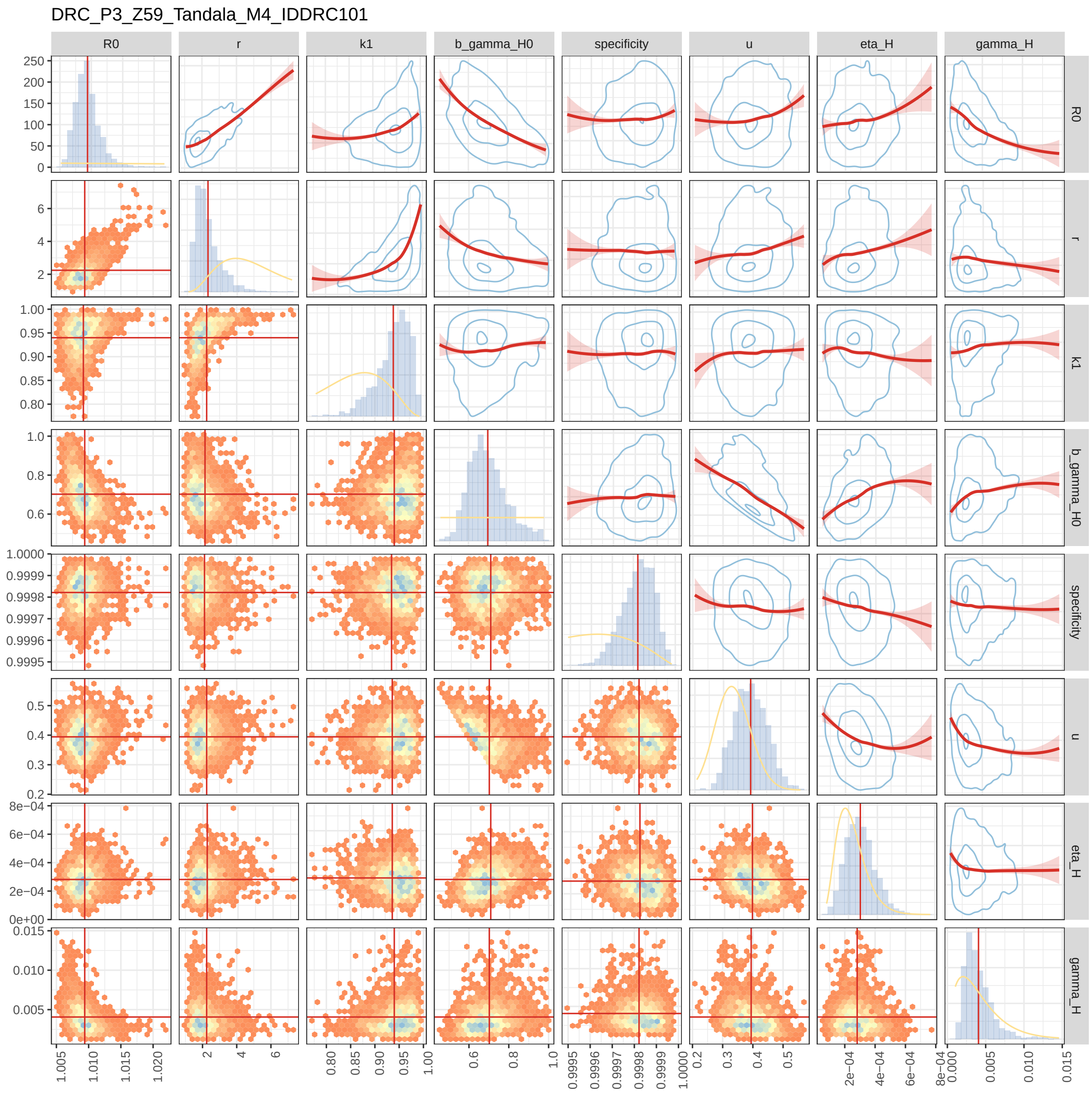


Figure S2.2: Joint posterior distributions for Tandala health zone in the former Equateur province.

#### S2.3 Posterior distribution maps

The posterior distribution maps (Figure S2.3–S2.16) illustrate both the level and variability of the parameter estimates between health zones. We partition the national or province level map into tessellated equilateral hexagons, or partial hexagons at national and health zone borders, and then fill each of these shapes with colour based on the value of randomly sampled values from the posterior distribution of the parameter being plotted from the analysis of the health zone in which that hexagon, or partial hexagon, lies. In this way, the 'overall' impression of the colour for a health zone gives an indication of the most probable value of the parameter, while the variation in colour reflects the uncertainty in the parameter estimates.

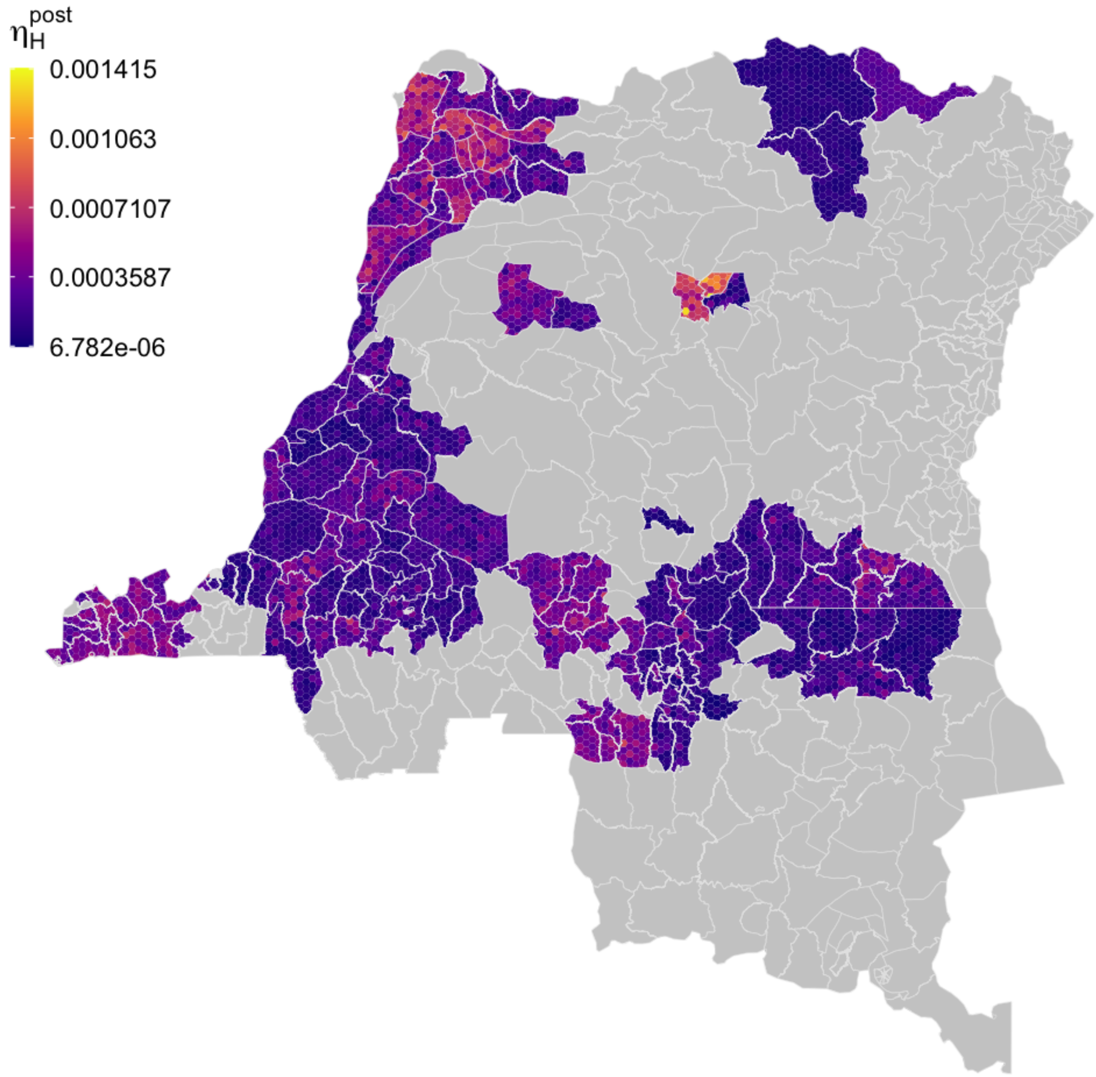


Figure S2.3: Within health zone posterior distribution of $\eta_H^{\text{post}}$, treatment rate from stage 1, 1998 onwards. fill colours are a randomly sampled from the posterior distribution for the health zone.

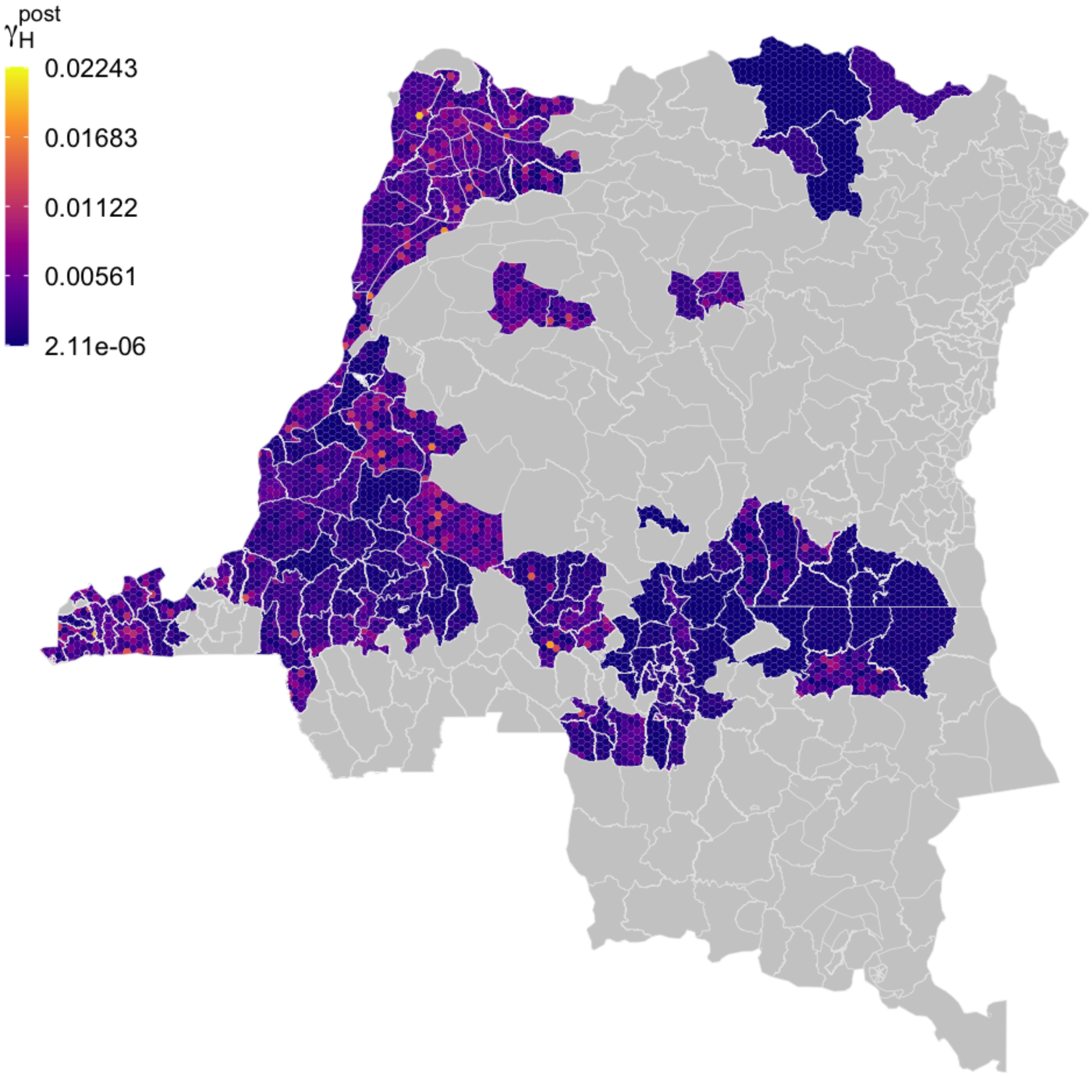


Figure S2.4: Within health zone posterior distribution of $\gamma_H^{\text{post}}$, the exit rate from stage 2 (treatment or death), 1998 onwards. The fill colours are a randomly sampled from the posterior distribution for the health zone.

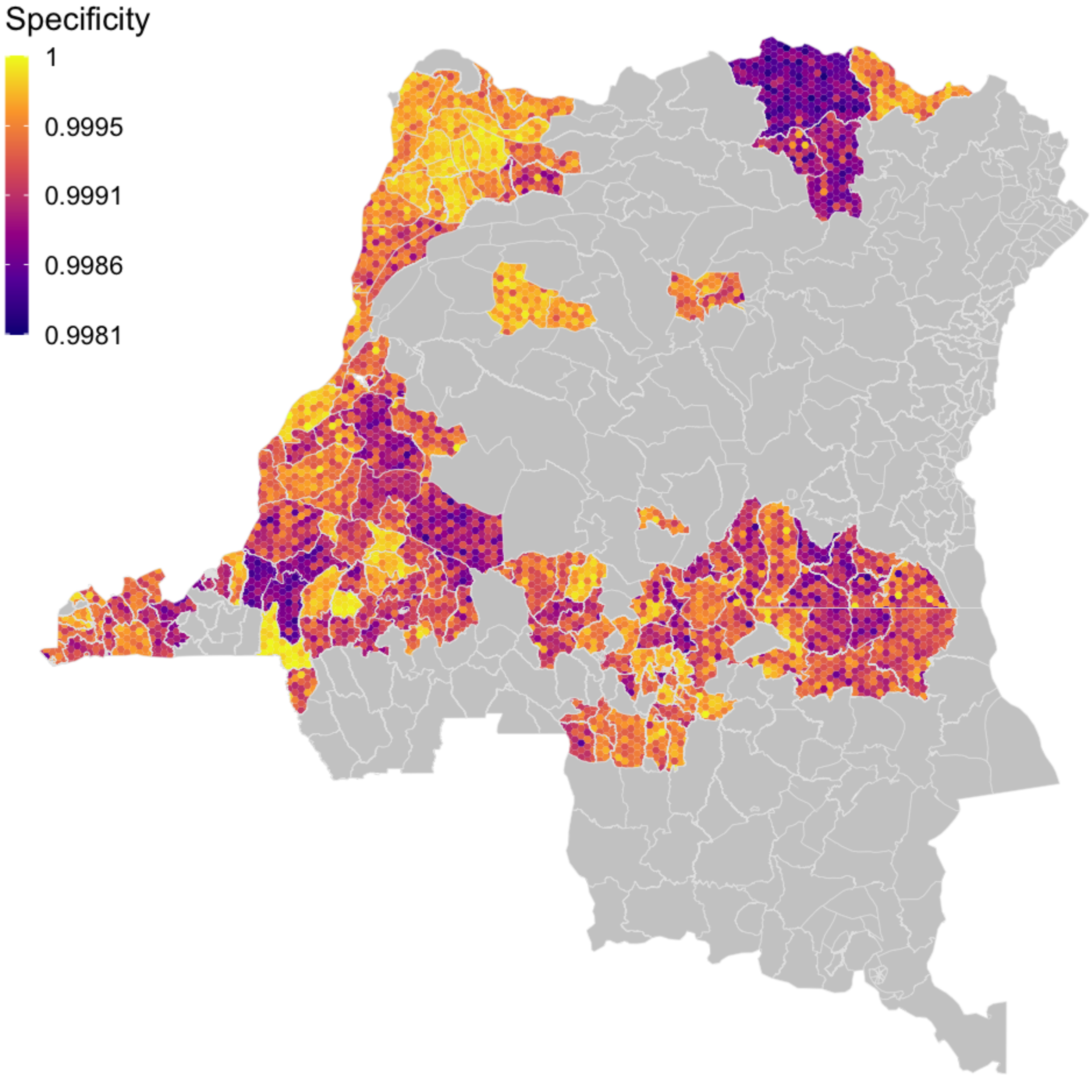


Figure S2.5: Within health zone posterior distribution of the specificity of the diagnostic algorithm, fill colours are a randomly sampled from the posterior distribution for the health zone.

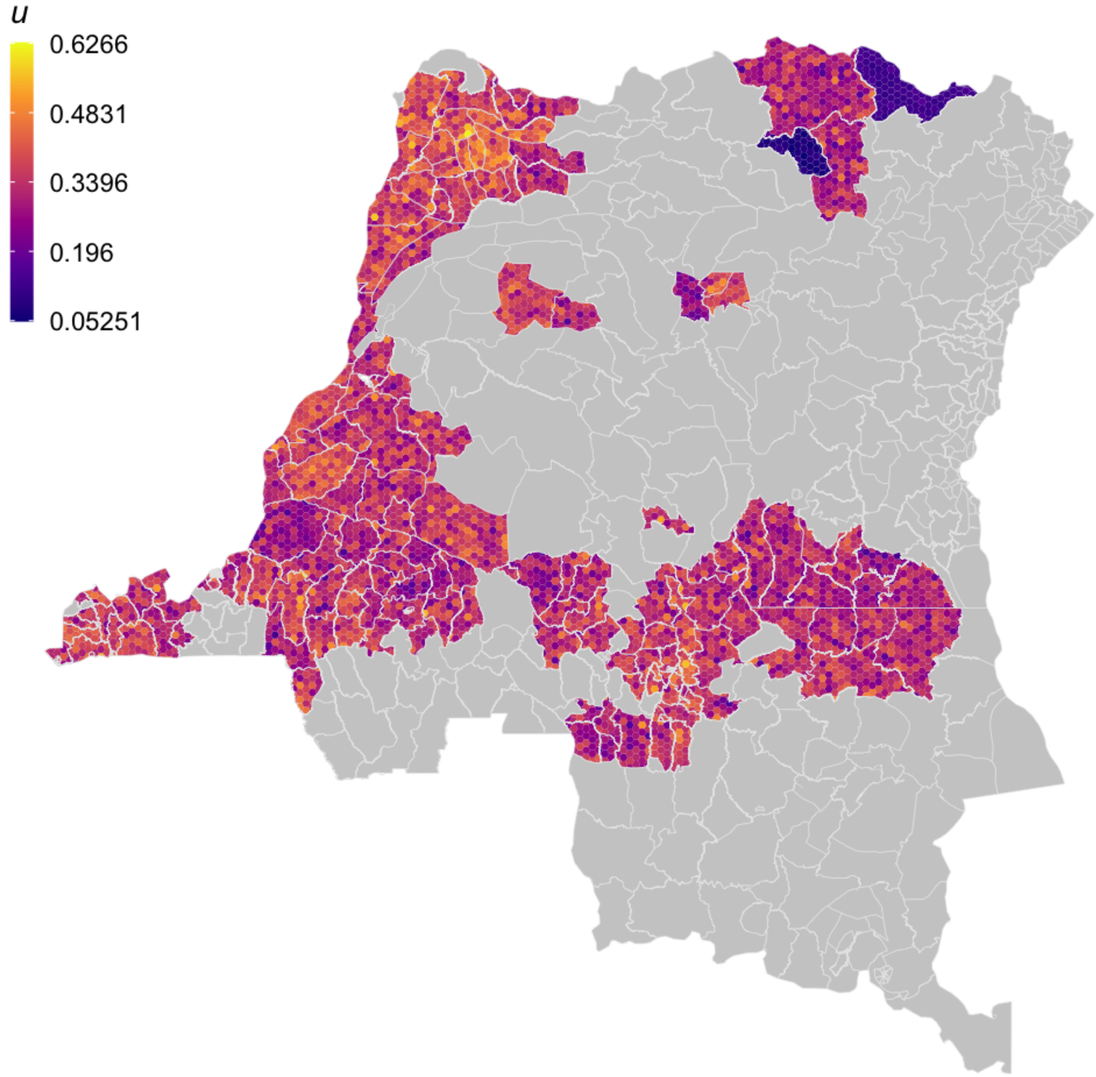


Figure S2.6: Within health zone posterior distribution of $u$, the proportion of stage 2 passive cases reported. The fill colours are a randomly sampled from the posterior distribution for the health zone.

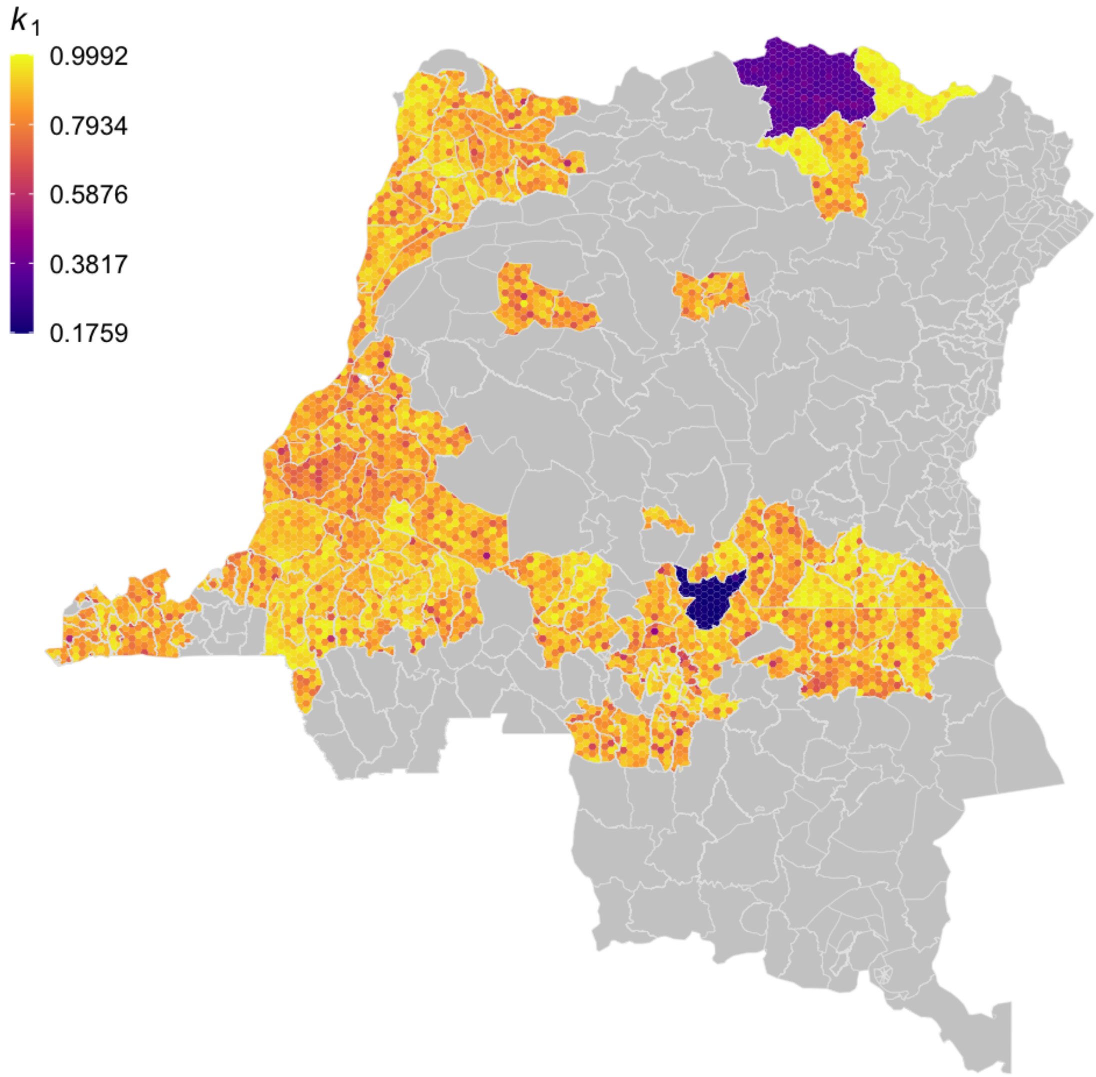


Figure S2.7: Within health zone posterior distribution of $k_1$, the proportion of low-risk people. The fill colours are randomly sampled from the posterior distribution for the health zone.

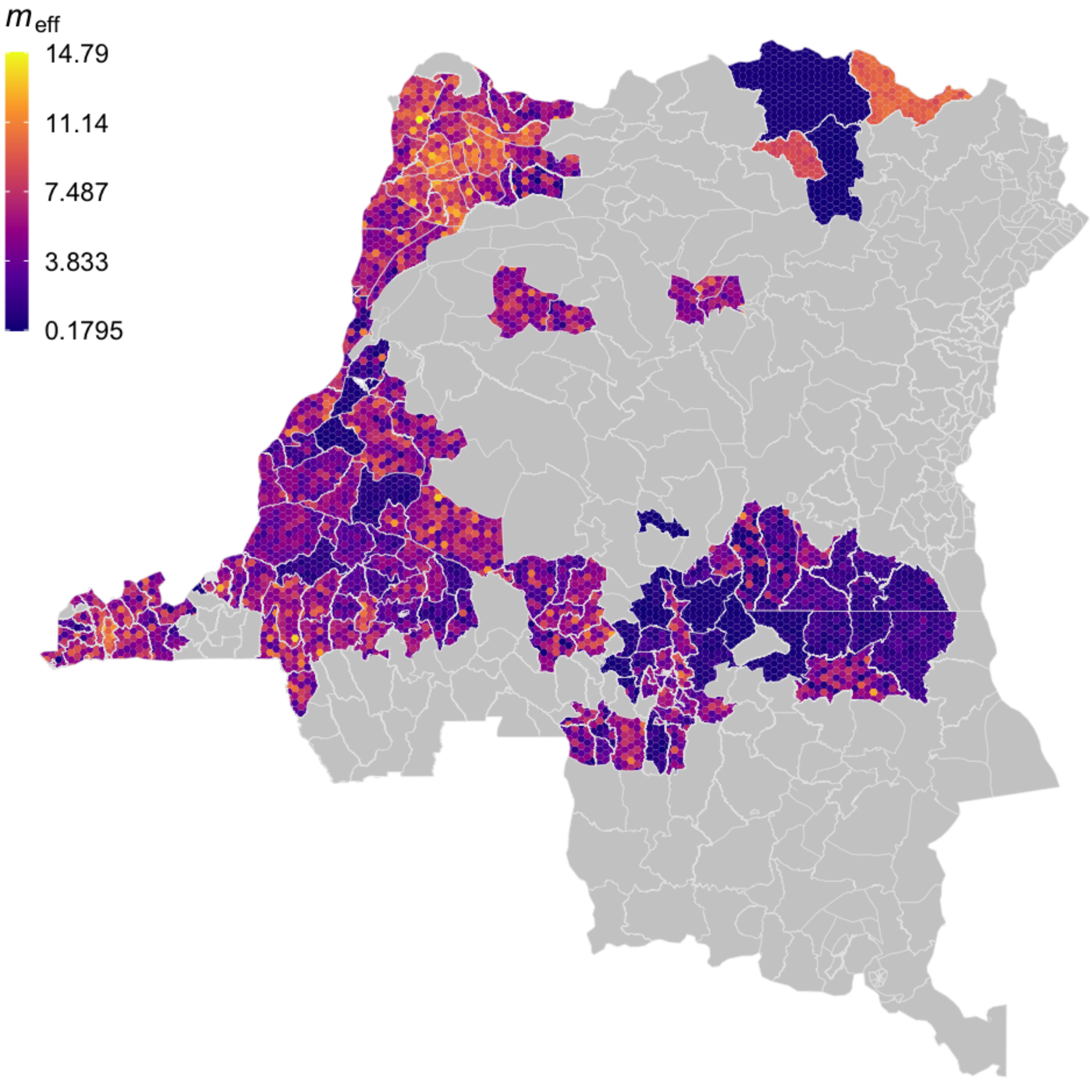


Figure S2.8: Within health zone posterior distribution of $m_{\mathrm{eff}}$; the effective, relative density of tsetse to humans. Fill colours are determined by randomly sampled values from the posterior distribution of $m_{\mathrm{eff}}$ from the analysis of the health zone.

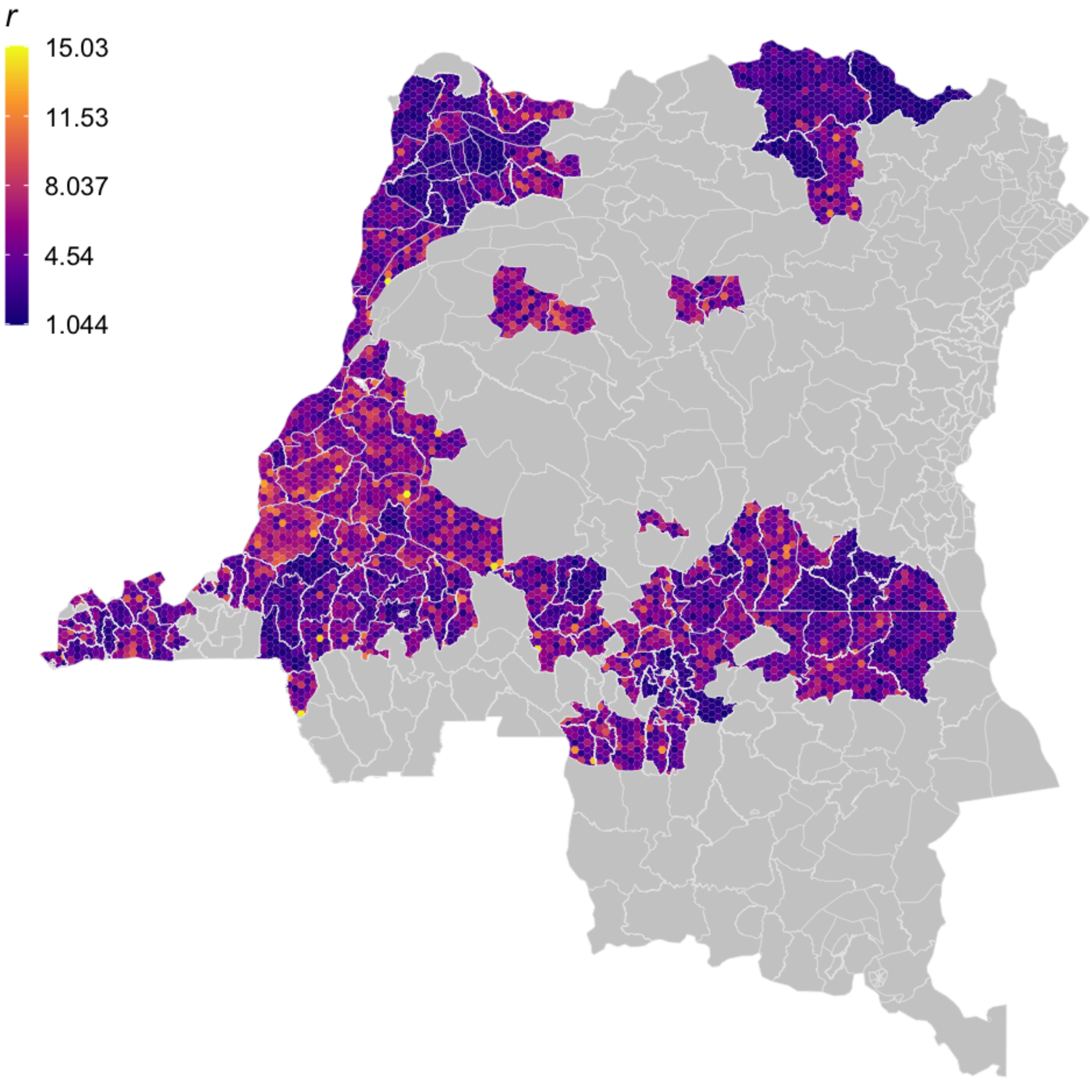


Figure S2.9: Within health zone posterior distribution of $r$, the relative bites taken on high-risk humans. Fill colours are determined by randomly sampled values from the posterior distribution of $r$ from the analysis of the health zone.

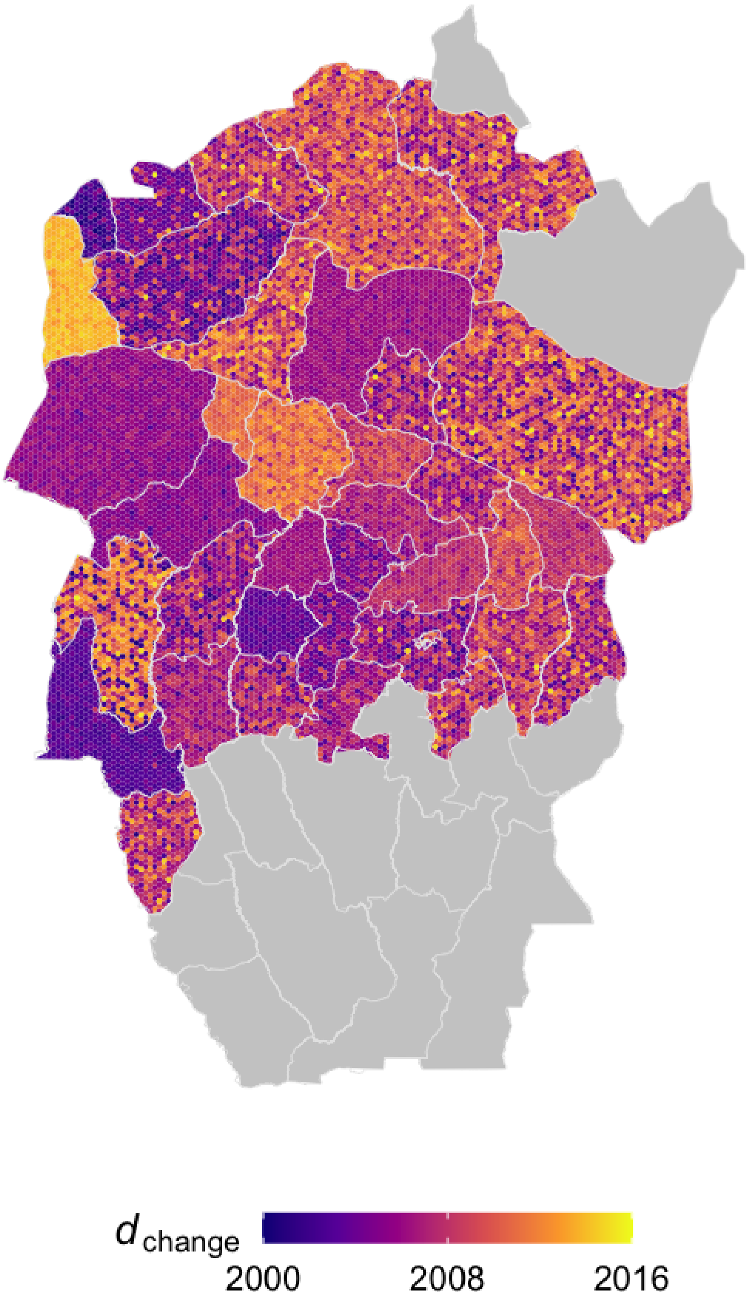


Figure S2.10: Within health zone posterior distribution of $d_{\text{change}}$, the midpoint year for passive improvement, for the former province of Bandundu. Fill colours are randomly sampled from the posterior distribution for the health zone.

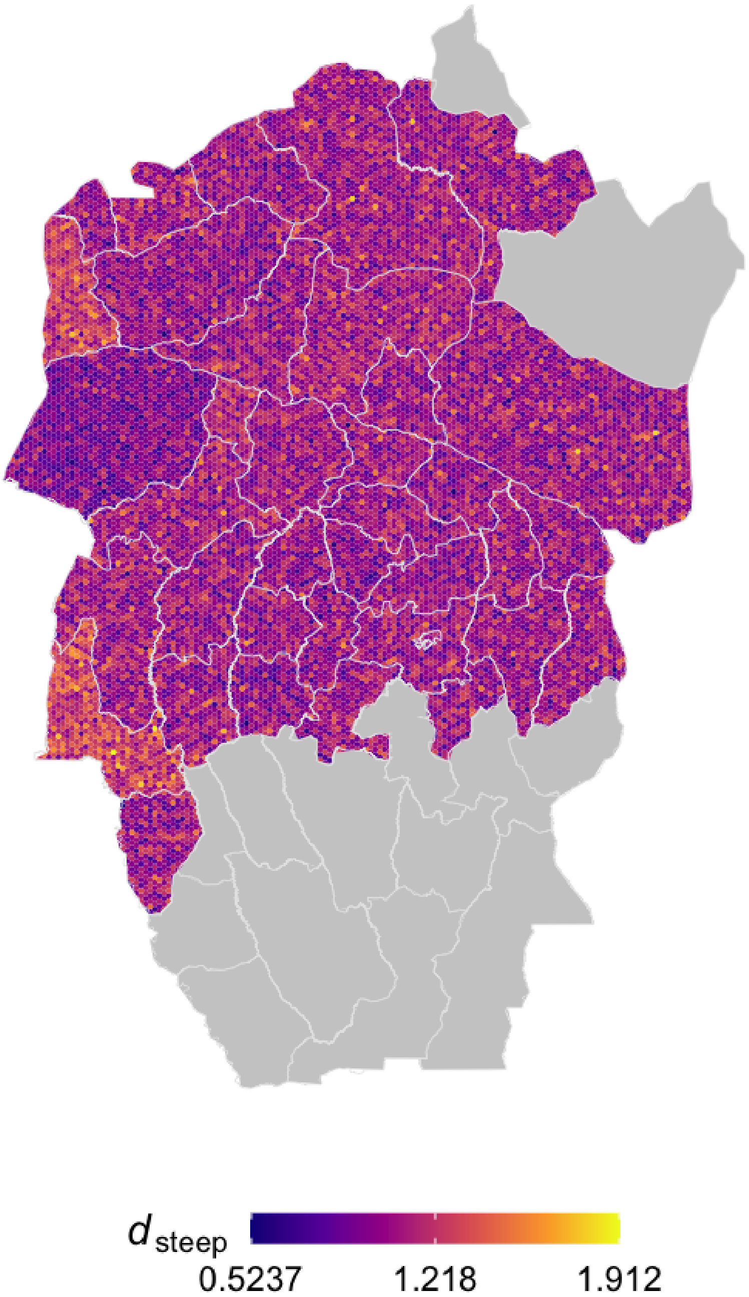


Figure S2.11: Within health zone posterior distribution of $d_{\text{steep}}$, the speed of improvement in passive detection rate, for the former province of Bandundu, fill colours are randomly sampled from the posterior distribution for the health zone.

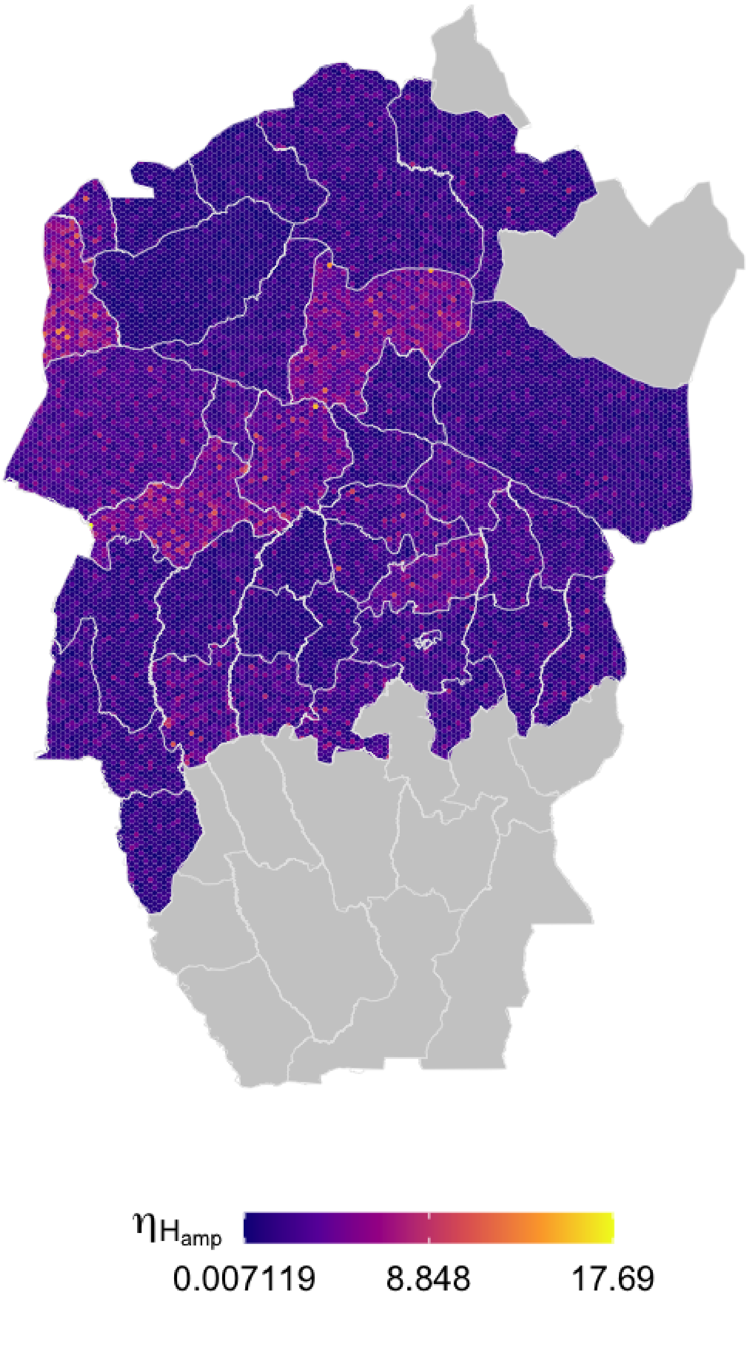


Figure S2.12: Within health zone posterior distribution of $\eta_{\mathrm{H}_{\mathrm{amp}}}$, the relative improvement in passive stage 1 detection rate, for the former province of Bandundu, fill colours are randomly sampled from the posterior distribution for the health zone.

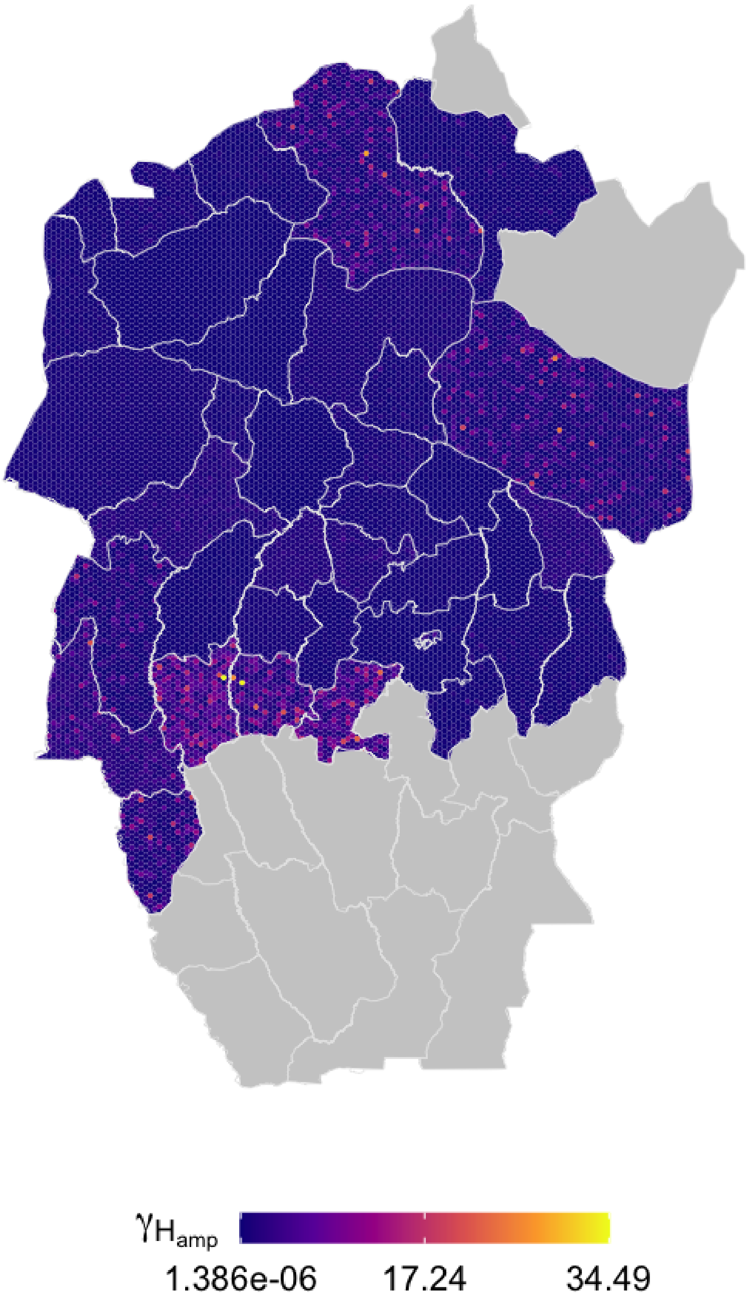


Figure S2.13: Within health zone posterior distribution of $\gamma_{\mathrm{H_{amp}}}$, the relative improvement in passive stage 2 detection rate, for the former province of Bandundu, fill colours are randomly sampled from the posterior distribution for the health zone.

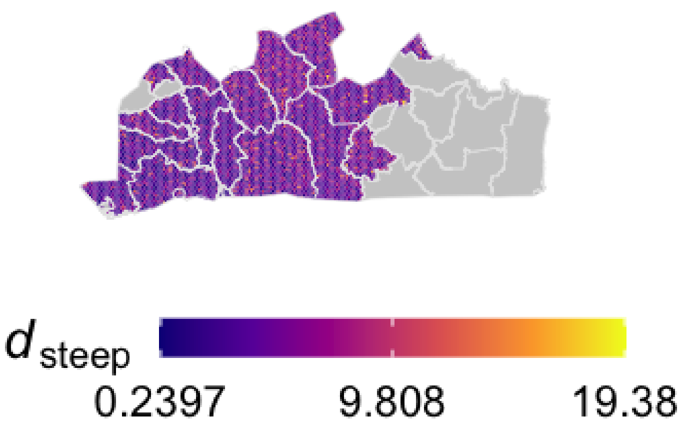


Figure S2.14: Within health zone posterior distribution of $d_{\text{steep}}$, the speed of improvement in passive detection rate, for the former province of Bas Congo, fill colours are randomly sampled from the posterior distribution for the health zone.

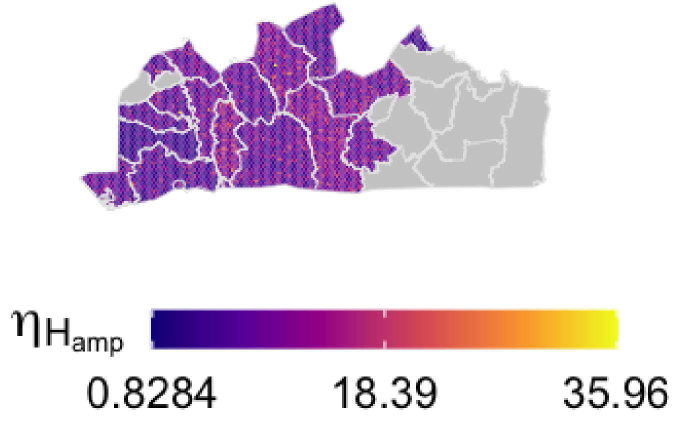


Figure S2.15: Within health zone posterior distribution of $\eta_{\text{H}_{\text{amp}}}$, the relative improvement in passive stage 1 detection rate, for the former province of Bas Congo, fill colours are randomly sampled from the posterior distribution for the health zone.

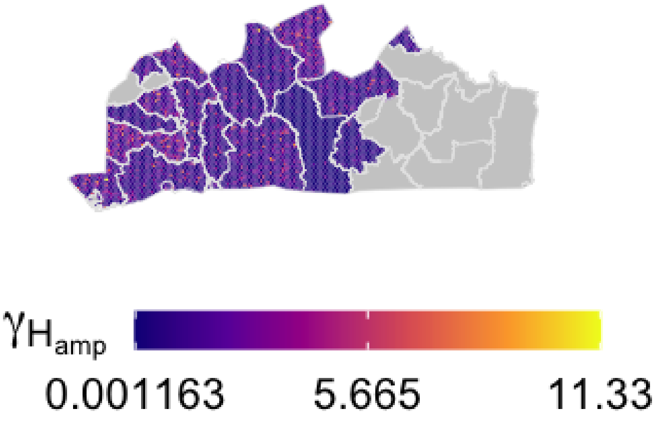


Figure S2.16: Within health zone posterior distribution of $\gamma_{H_{amp}}$, the relative improvement in passive stage 2 detection rate, for the former province of Bas Congo, fill colours are randomly sampled from the posterior distribution for the health zone.
