## Supplementary material for "Quantifying epidemiological drivers of *gambiense* human African Trypanosomiasis across the Democratic Republic of Congo": Information on inference of time infected and case reporting from parameter samples

### Supplementary Information: Quantifying epidemiological drivers of *gambiense* human African trypanosomiasis across the Democratic Republic of Congo. S3 Using posteriors to infer time infected and reporting

Ronald E Crump[1,2,3◑*], Ching-I Huang[1,2◑], Ed Knock[1,4], Simon E F Spencer[1,4], Paul Brown[1,2], Erick Mwamba Miaka[5], Chansy Shampa[5], Matt J Keeling[1,2,3], and Kat S Rock[1,2]

[1]Zeeman Institute for System Biology and Infectious Disease Epidemiology Research, The University of Warwick, Coventry, U.K.
[2]Mathematics Institute, The University of Warwick, Coventry, U.K.
[3]The School of Life Sciences, The University of Warwick, Coventry, U.K.
[4]The Department of Statistics, The University of Warwick, Coventry, U.K.
[5]PNLTHA, Kinshasa, D.R.C.

October 26, 2020

◑These authors contributed equally to this work.

#### S3.1 Time spent infected

In order to provide a straightforward metric to assess the improvements to passive detection over time we compute the average time spent infected by people not picked up by active screening (for example by people in the high-risk group) using the following equation:

$$\begin{aligned} T_{\text{infected}}(Y) &= \mathbb{P}(\text{Passively detected in S1}) \\ &\qquad \times \text{Time spend infected in S1} \\ &\quad + \mathbb{P}(\text{Passively detected in S2 or unreported}) \\ &\qquad \times \text{Time spend infected in S1 and S2} \\ &= \left[\frac{\eta_H(Y)}{\eta_H(Y)+\varphi_H} \times \frac{1}{\eta_H(Y)+\varphi_H}\right] \\ &\quad + \left[\frac{\varphi_H}{\eta_H(Y)+\varphi_H} \times \left(\frac{1}{\gamma_H(Y)} + \frac{1}{\eta_H(Y)+\varphi_H}\right)\right] \end{aligned} \tag{S3.1.1}$$

The inferred change in average time spent infected in Kwamouth and Tandala is shown in Figure S3.1; in Kwamouth health zone the time has decreased due to improvements in passive surveillance. In Kwamouth the average time changes from 1224 (95% CI: 690–2547) days to 813 (95% CI: 548–1349) days. In Tandala no improvement in passive detection over time was modelled, and hence the amount of time spent infected remains at 716 days on average (95% CI: 548–974). Active screening will have further brought these durations down for those in the population who participate in active screening (low-risk individuals). It is interesting to note that our estimates for mean time spent infected are a little larger than those estimated by Checchi *et al* [S1], even though their estimate for stage 2 duration informed our prior on $\gamma_H^{\text{post}}$; their combined estimate of expected duration (S1 and S2 with no treatment) is 778 days (95% CI: 525–1232).

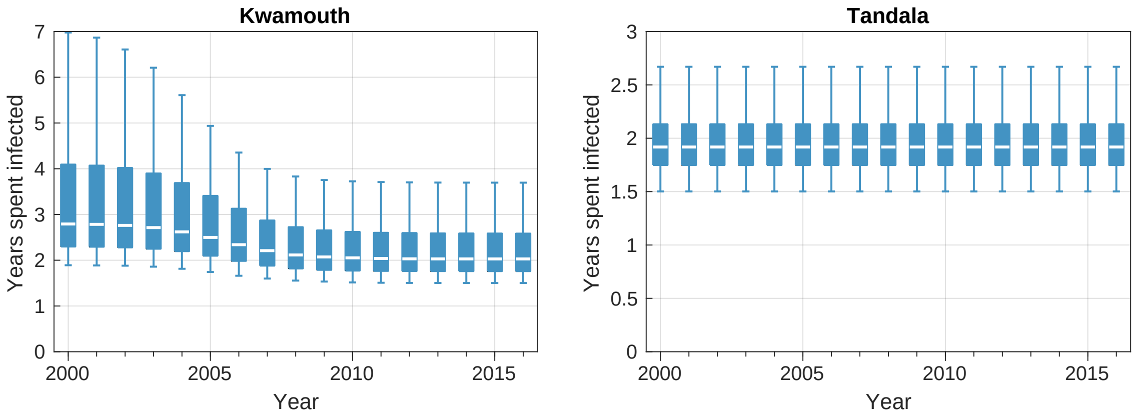


Figure S3.1: Change in the average time spend infected if not picked up by active screening. Kwamouth is shown on the left, and Tandala on the right.

#### S3.2 Proportion of successful treatments or deaths reported

In order to estimate the change in the proportion of infections reported over time we use sampled model outputs of deaths and case reporting (active and passive for both stages):

$$\text{Proportion reported} = 1 - \frac{\text{Deaths}}{\text{Active1} + \text{Active2} + \text{Passive1} + \text{Passive2} + \text{Deaths}} \qquad \text{(S3.2.1)}$$

The estimated change in proportion reported over time in Kwamouth and Tandala is shown in Figure S3.2; in both locations the proportion of reporting is inferred to vary in time, although the overall decrease in infection and case reporting results in larger credible intervals for 2016, especially in Tandala. In Kwamouth the model estimates the proportion of cases and deaths reported changed from 0.67 (95%: 0.54–0.84) in 2000 to 0.82 (95%: 0.62–0.93) in 2016; the consistent high level active screening in conjunction with improving passive surveillance results in an improving trend through time. In Tandala the proportion reported fluctuated over time from 0.65 (95%: 0.53–0.75) in 2000 and finishing back at 0.65 (95%: 0.0–1.0) in 2016 with the median varying between 0.40 (in 2010) and 0.76 (in 2003); the amount of active screening was substantially lower in 2008–2015 than in the 2000–2007 period which is why the average proportion reported is lower in that time period.

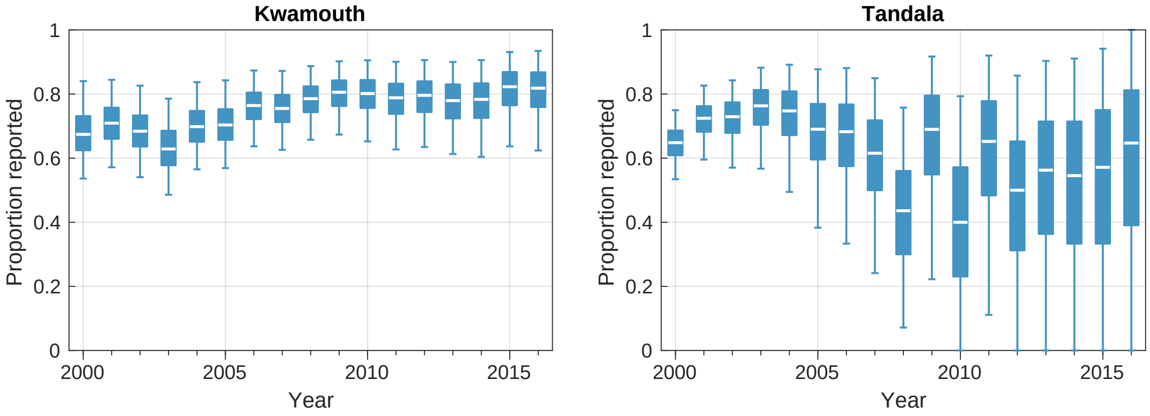


Figure S3.2: Change in the estimated proportion of infections reported over time. Box plots show estimates for the median (center line), 50% percentiles (boxes) and 95% percentiles (whiskers). Kwamouth is shown on the left, and Tandala on the right.
