## Supplementary material for "Quantifying epidemiological drivers of *gambiense* human African Trypanosomiasis across the Democratic Republic of Congo": Addressing the PRIME-NTD criteria for good modelling practises

### Supplementary Information: Quantifying epidemiological drivers of *gambiense* human African trypanosomiasis across the Democratic Republic of Congo. S4 PRIME-NTD criteria

Ronald E Crump[1,2,3◑*], Ching-I Huang[1,2◑], Ed Knock[1,4], Simon E F Spencer[1,4], Paul Brown[1,2], Erick Mwamba Miaka[5], Chansy Shampa[5], Matt J Keeling[1,2,3], and Kat S Rock[1,2]

[1]Zeeman Institute for System Biology and Infectious Disease Epidemiology Research, The University of Warwick, Coventry, U.K.
[2]Mathematics Institute, The University of Warwick, Coventry, U.K.
[3]The School of Life Sciences, The University of Warwick, Coventry, U.K.
[4]The Department of Statistics, The University of Warwick, Coventry, U.K.
[5]PNLTHA, Kinshasa, D.R.C.

October 26, 2020

◑These authors contributed equally to this work.

#### S4.1 PRIME-NTD criteria

It has been recommended that good modelling practises should meet the five key principles relating to communication, quality and relevance of analyses - known as Policy-Relevant Items for Reporting Models in Epidemiology of Neglected Tropical Diseases (PRIME-NTD) [S1]. We demonstrate how these PRIME-NTD criteria have each been addressed in Table S4.1.

Table S4.1: PRIME-NTD criteria fulfillment. How the NTD Modelling Consortium's "5 key principles of good modelling practice" have been met in the present study.

| Principle | What has been done to satisfy the principle? | Where in the manuscript is this described? |
|---|---|---|
| **1. Stakeholder engagement** | | |
| | This study was lead by modellers and guided by members of the national sleeping sickness control programme in DRC (PNLTHA-DRC) – coauthors E Mwamba Miaka and S Chancy. PNLTHA-DRC have contributed to improved modeller understanding of the epidemiological data and changes to the programme over time and in different geographic regions, both of which impacted model fitting over several rounds of revision (via in-person meetings and email). The GUI (and several variants of it) was designed in conjunction with PNLTHA-DRC to improve communication of the modelling outputs to non-modellers. It has been refined through various in-person meetings with different collaborators with the goal of providing understandable, policy-relevant outputs as well as scientific communication; over 20 non-modellers have had opportunities to interact with and provide feedback on the GUI during development. | Authorship list |
| **2. Complete model documentation** | | |
| | Full model fitting code and documentation is available through OpenScienceFramework (OSF). The model is fully described in the main text and SI. | See Materials and Methods section in the main text, Supplementary Information (file S1) and at OSF (`https://doi.org/10.17605/osf.io/ck3tr`) |
| **3. Complete description of data used** | | |
| | The original data and how we aggregated the data for fitting were described in detail in the main text and SI. Aggregate data can be viewed next to model fits in our GUI. | See Materials and Methods section, Supplementary Information (file S1, section S1.1) and the GUI (`https://hatmepp.warwick.ac.uk/fitting/v2/`) |
| **4. Communicating uncertainty** | | |
| | *Structural uncertainty*: The variant of the model presented here ("Model 4") was chosen as it had good support compared to other plausible model structures when fitting to data sets from Yasa Bonga and Mosango health zones in DRC [S3] and in the Mandoul focus, Chad [S2]. | *Structural uncertainty*: Materials and Methods section in main text. |
| | *Parameter uncertainty*: We provide estimates for the parameter uncertainty in each health zone within our posterior parameter maps (randomly sampled values from posteriors) and joint posterior distributions of fitted parameters (main text and SI) and also show distributions (histograms) in the GUI. | *Parameter uncertainty*: Figure 6, Supplementary Information (S2) Figures S2.1–S2.16 and model uncertainty maps in GUI (`https://hatmepp.warwick.ac.uk/fitting/v2/`) |

Continued on next page

**Table S4.1 – continued from previous page**

| Principle | What has been done to satisfy the principle? | Where in the manuscript is this described? |
|---|---|---|
| | *Prediction uncertainty*: We represent uncertainty in our results by: (i) summarising province-level estimates of changes to new infections; and (ii) providing box and whisker plots for fitted dynamics (median, 50% and 95% credible intervals). | *Prediction uncertainty*: (i) Table 3 (ii) Figure 5 |
| **5. Testable model outcomes** | | |
| | Previous versions of this model have undergone validation exercises (data censoring) to examine the robustness of the predictive ability of the model [S2, S4]. Whilst this was not performed here, the multiple rounds of model fitting, critical review and refinement in discussion with PNLTHA produced very clear improvements to the fit by altering assumptions about the passive detection rates and changes to diagnostic specificity over time. This was most improved for former Bandundu province where we are now able to match "humped" trends in passive detection. | See main text results for explanation of "humped" trends in passive reporting. |
